# Plasma neurofilament light chain predicts Alzheimer’s disease in patients with subjective cognitive decline and mild cognitive impairment: a longitudinal study

**DOI:** 10.1101/2023.05.19.23290183

**Authors:** Salvatore Mazzeo, Silvia Bagnoli, Assunta Ingannato, Sonia Padiglioni, Giulia Giacomucci, Alberto Manganelli, Valentina Moschini, Juri Balestrini, Arianna Cavaliere, Carmen Morinelli, Giulia Galdo, Filippo Emiliani, Diletta Piazzesi, Chiara Crucitti, Daniele Frigerio, Cristina Polito, Valentina Berti, Sandro Sorbi, Benedetta Nacmias, Valentina Bessi

## Abstract

**Background:** We aimed to evaluate the accuracy of plasma neurofilament light chain (NfL) in predicting Alzheimer’s disease (AD) and the progression of cognitive decline in patients with subjective cognitive decline (SCD) and mild cognitive impairment (MCI).

**Methods:** This longitudinal cohort study involved 140 patients (50 with SCD, 73 with MCI, and 22 with AD dementia [AD-D]) who underwent plasma NfL and AD biomarker assessments (CSF, amyloid-PET, and ^18^F-FDG-PET) at baseline. They were rated according to the A/T/N system and followed up for a mean time of 2.72±0.95 years to detect progression from SCD to MCI and from MCI to AD. Forty-eight patients (19 SCD, 29 MCI) also underwent plasma NfL measurements after two years after baseline.

**Results:** At baseline, plasma NfL detected patients with biomarker profiles consistent with AD (A+/T+/N+ or A+/T+/N-) with high accuracy (AUC=0.82). We identified cut-off value of

19.45 pg/mL for SCD and 20.45 pg/mL for MCI. During follow-up, nine SCD patients progressed to MCI (p-SCD), and 14 MCI patients developed AD dementia (p-MCI). The previously identified cut-off values provided good accuracy in identifying p-SCD (80% [95% C.I.=65.69:94.31]). The rate of NfL change was higher in p-MCI (3.52±4.06 pg/mL) compared to np-SCD (0.81±1.25 pg/mL) and np-MCI (−0.13±3.24 pg/mL) patients. A rate of change lower than 1.64 pg/mL per year accurately excluded progression from MCI to AD (AUC=0.954).

**Conclusion:** Plasma NfL concentration and change over time may be a reliable, non-invasive tool to detect AD and the progression of cognitive decline at the earliest stages of the disease.

**Key messages:**

- **What is already known on this topic** Plasma NfL increase in SCD, MCI and AD and longitudinal changes in NfL are related to changes in brain atrophy and cognitive outcomes in AD. Nevertheless, the clinical value of plasma NfL in non-demented patients has been poorly explored.
- **What this study adds** Plasma NfL accurately predicts AD pathology and progression of cognitive decline in SCD and MCI. Repeated measurements of NfL may further increase the accuracy of this biomarker
- **How this study might affect research, practice, or policy** Given its accessibility, blood-based NfL can assist clinicians in determining the optimal personalized diagnostic and therapeutic approach for individuals presenting with SCD or MCI, providing insights into the underlying biological mechanisms of cognitive decline, even in primary care settings.

## 1. INTRODUCTION

Subjective cognitive decline (SCD)^1^ and mild cognitive impairment (MCI)^2^ are considered as first presentations of Alzheimer’s disease (AD)^3^ and represent the main target population for selecting patients for clinical trials and for the administration of upcoming disease modifying therapies (DMTs)^4^. Nevertheless, MCI and SCD are very common and heterogeneous conditions with several possible trajectories and many potential underlying causes^5^. Neurofilament light chain (NfL), a component of the neuronal cytoskeleton^6^, has recently emerged as a promising blood-based biomarker for AD^7, 8^. Elevated levels of plasma NfL have been observed in individuals with SCD^9^, MCI^10^ and dementia due to AD (AD-D)^11^ compared to cognitively normal individuals and longitudinal changes in NfL are related to changes in brain atrophy and cognitive outcomes in AD^11^. Nevertheless, the accuracy of this biomarker in predicting a possible underlying AD pathology^12, 13^ and the clinical meaning of NfL level change over time in non-demented patients^14, 15^ has been poorly explored so far. In this perspective, we hypothesized that plasma NfL level and its change over time may mirror the underlying AD biomarker profile and predict the progression of cognitive decline.

## 2. MATERIALS AND METHODS

### 2.1. Patients

We enrolled 140 consecutive patients (45 SCD, 73 MCI, 22 AD-D) referred to the Centre for Adult Cognitive Disorders of Careggi Hospital in Florence for assessment of cognitive decline, between July 2018 and November 2022. We included patients who met criteria for clinical diagnosis of AD-D^16^, MCI^2^, or SCD^1^. Exclusion criteria were: history of head injury, current systemic and/or neurological disease other than AD, major depression or substance use disorder. At baseline, all patients underwent comprehensive clinical assessment, neurological examination, extensive neuropsychological investigation (as described in detail elsewhere^17^), and blood collection for measurement of plasma NfL concentration and Apolipoprotein E (*APOE)* genotype analysis. We defined age at baseline as the age at the time of plasma collection, disease duration as the time from the onset of symptoms to baseline examination, and positive family history of dementia if one or more first-degree relatives were reported to have documented cognitive decline. One-hundred-ten patients (30 SCD, 60 MCI, 20 AD-D) underwent CSF collection for Aβ_42_, Aβ_42_/Aβ_40_, total-tau (t-tau) and phosphorylated-tau (p-tau). Among these, 28 patients (16 SCD, 9 MCI, 3 AD-D) also underwent cerebral amyloid-PET, and 93 patients (23 SCD, 51 MCI, 19 AD-D) also underwent ^18^F-Fluorodeoxyglucose-PET brain scan (FDG-PET). Normal values for CSF biomarkers were: Aβ_42_>670 pg/ml, Aβ_42_/Aβ_40_ ratio>0.062, t-tau<400 pg/ml and p-tau<60 pg/ml^18^.

Methods used for *APOE* genotyping^19^, CSF collection and biomarker analysis, brain ^18^F-FDG-PET and amyloid-PET acquisition and rating^20^ are described in further detail elsewhere (see Bessi et al.^19^ and Mazzeo et al.^20^ respectively).

Seventy-seven patients (30 SCD and 47 MCI) underwent neuropsychological examination after two years. From 48 of them (19 SCD, 29 MCI) blood samples were collected two years after baseline to repeat the NfL measure. Progression to MCI and to AD was defined according to the National Institute on Aging-Alzheimer’s Association (NIA-AA) criteria^2, 16^ by two trained neurologists (SM and VB) who were blinded to the plasma NfL results.

### 2.2. Standard Protocol Approvals

The study procedures and data analysis were performed in accordance with the Declaration of Helsinki and with the ethical standards of the Committee on Human Experimentation of our Institute. This study was approved by the Institutional Review Board of Careggi University Hospital (Florence, Italy, reference 15691oss). All individuals involved in this research agreed to participate and to have details and results of the research about them published.

### 2.3. Plasma collection and NfL analysis

Blood was collected by venipuncture into standard polypropylene EDTA test tubes (Sarstedt, Nümbrecht, Germany) and centrifuged within two hours at 1300 rcf at 4°C for 10 minutes. Plasma was isolated and stored at −80°C until testing. Plasma NfL analysis was performed with Simoa NF-Light SR-X kit (cat. No. 103400) for human samples provided by Quanterix Corporation (Lexington, MA, USA) on the automatized Simoa SR-X platform (GBIO, Hangzhou, China), following the manufacturer’s instructions. The lower limits of quantification and detection provided by the kit were 0.316 and 0.0552 pg/mL, respectively. The plasma NfL concentrations in all samples were detected in a single run. Quality controls with a low NfL concentration of 5.08 pg/mL and a high NfL concentration of 169 pg/mL were included in the array and assessed with samples. A calibration curve was constructed from the measurements of serially diluted calibrators provided by Quanterix. Plasma samples and controls were diluted at a 1:4 ratio and measured in duplicate with calibrators.

### 2.4. Classification of patients according to the ATN classification

Based on biomarker results, patients were classified according to the NIA-AA Research Framework^21^: patients were rated as A+ if at least one of the amyloid biomarkers (CSF or amyloid PET) revealed the presence of Aβ pathology, and as A- if none of the biomarkers revealed the presence of Aβ pathology. In the case of discordant CSF and Amyloid PET results, we considered only the pathological result. Patients were classified as T+ or T- if CSF p-tau concentrations were higher or lower than the cut-off value, respectively. Patients were classified as N+ if at least one neurodegeneration biomarker was positive (CSF t-tau higher than the cut-off value or positive FDG-PET). Considering our sample size, to avoid too small groups, we considered T and N parameters together as TN+ if they were T+ and/or N+, and TN- if both T and N were negative. Finally, we defined four groups: normal AD biomarkers (A-/T-/N-), non-AD pathologic change (A-/TN+), Alzheimer’s pathologic change (A+, including A+/T-/N-patients and one patient with A+/T-/N+), and AD (A+/T+/N+, including A+/TN+ and A+/T+/N+).

### 2.5. Statistical analysis

All statistical analyses were performed using IBM SPSS Statistics Software Version 25 (SPSS Inc., Chicago, USA) and the computing environment R4.2.3 (R Foundation for Statistical Computing, Vienna, 2013). All p-values were two-tailed and the significance level for all analyses was set at p=0.05. Distributions of all variables were assessed using the Shapiro-Wilk test. As NfL was not normally distributed, we applied log10 transformation. This transformation resulted in a more normally distributed dataset that met the assumptions of the statistical tests that we planned to use. We conducted descriptive statistics using means and standard deviations (SD) for continuous variables and frequencies or percentages and 95% confidence intervals (95%C.I.) for categorical variables. We used t-test for comparison between two groups, one-way analysis of variance (ANOVA) with Bonferroni post-hoc test for comparison between three or more groups, Pearson’s correlation coefficient to evaluate correlations between groups’ numeric measures, and chi-square test to compare categorical data. To adjust for possible confounding factors, we used analysis of covariance (ANCOVA). We constructed receiver operating characteristic (ROC) curves to evaluate the performance of plasma NfL in predicting ATN status and progression of cognitive decline. We used the Youden method to determine the optimal cut-off value for NfL and calculated accuracy, sensitivity, specificity, positive predictive value (PPV), and negative predictive value (NPV). We used Kaplan-Meier survival analyses with pairwise log-rank to compare proportions of progression of cognitive decline among groups. We used Cox regression analysis to ascertain that the effect of NfL on progression from SCD to MCI was independent from other covariates. The consistency of NfL measures over time was computed using the intraclass correlation coefficient (ICC). We used repeated measures ANOVA to investigate the effect of progression of cognitive decline and ATN status on the change in NfL concentration over time. We calculated the size effect using Cohen’s *d* for normally distributed numeric measures, η^2^ for ANOVA and Cramer’s *V* for categorical data.

## 3. RESULTS

### 3.1. Comparisons between groups

NfL levels were significantly different between SCD, MCI and AD-D groups (*F*[2,136]=14.99, *p*<0.001, η^2^=0.181) (Figure 1.A). Demographic features and differences between groups are summarized in Table 1. NfL concentration was correlated with age at baseline (Pearson=0.549, p<0.001) and MMSE (Pearson=-0.291, p=0.001). There were no differences in NfL concentrations between males and females (p=0.222) or between *APOE*ε4+ and *APOE*ε4-(p=0.579). The significant effect of diagnosis group (SCD, MCI and AD-D) on NfL concentration was confirmed after controlling for age, education, MMSE and *APOE* genotype (*F*[2,119]=3.51, p=0.033, partial η^2^=0.056, Supplementary table 1.A).

**Figure 1.**
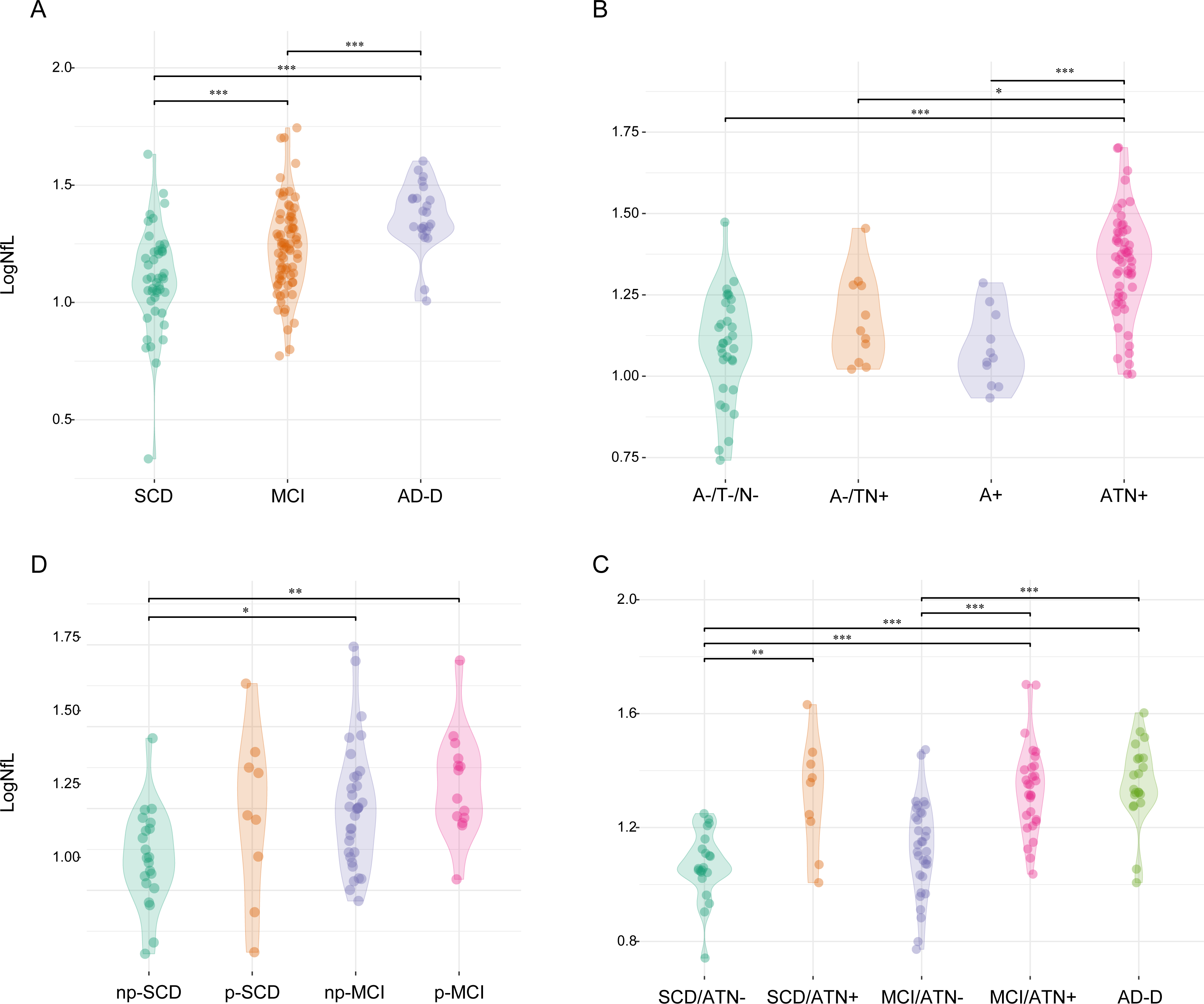
LogNfL levels across groups. Values quoted in the y-axis indicate LogNfL levels. Horizontal bars indicate significant differences between groups. A. Comparisons between diagnosis groups: SCD vs. MCI (p = 0.002, Cohen’s d = 0.671); SCD vs. AD (p <□0.001, Cohen’s d = 1.394); MCI vs. AD (p = 0.010, Cohen’s d = 0.723). B. Comparisons between ATN groups: A-/T-/N- vs. A-/TN+ (p = 0.765, Cohen’s d = 0.537); A-/T-/N- vs. A+/T-/N- (p = 1.00, Cohen’s d = 1.249); A-/T-/N- vs. A+ (p < 0.001, Cohen’s d = 1.562); A-/T-/N- vs. ATN+ (p < 0.001, Cohen’s d = 1.562); A-/TN+ vs. A+ (p = 1.00, Cohen’s d = 0.394); A-/TN+ vs. ATN+ (p < 0.001, Cohen’s d = 1.419); A+ vs. ATN+ (p < 0.01, Cohen’s d = 1.419) C. Comparisons diagnosis/ATN groups: SCD/ATN- vs. SCD/ATN+ (p = 0.003, Cohen’s d = 1.571, MCI/ATN+ (p <□0.001, Cohen’s d = 1.747); SCD/ATN- vs. AD (p <□0.001, Cohen’s d = 1.880); MCI/ATN- vs. MCI/ATN+ (p <□0.001, Cohen’s d = 1.343); MCI/ATN- vs. AD (p <□0.001, Cohen’s d = 1.476); SCD/ATN- vs. MCI/ATN- (p = 1.00, Cohen’s d = 0,447); SCD/ATN+ vs. MCI/ATN+ (p = 1.00, Cohen’s d = 0.176); MCI/ATN+ vs. AD (p = 1.00, Cohen’s d = 0.133). D. Comparisons between progression groups: np-SCD vs. p-SCD (p = 0.428, Cohen’s d = 0.729); np-SCD vs. np-MCI (p = 0.020, Cohen’s d = 0.846); np-SCD vs. p-MCI groups (p = 0.003, Cohen’s d = 1.250); p-SCD vs. np-MCI (p = 1.00, Cohen’s d = 0.117), p-SCD vs. p-MCI (p = 1.00, Cohen’s d = 0.521); np-MCI vs. p-MCI (p = 1.00, Cohen’s d = 0.404).

**Table 1.**
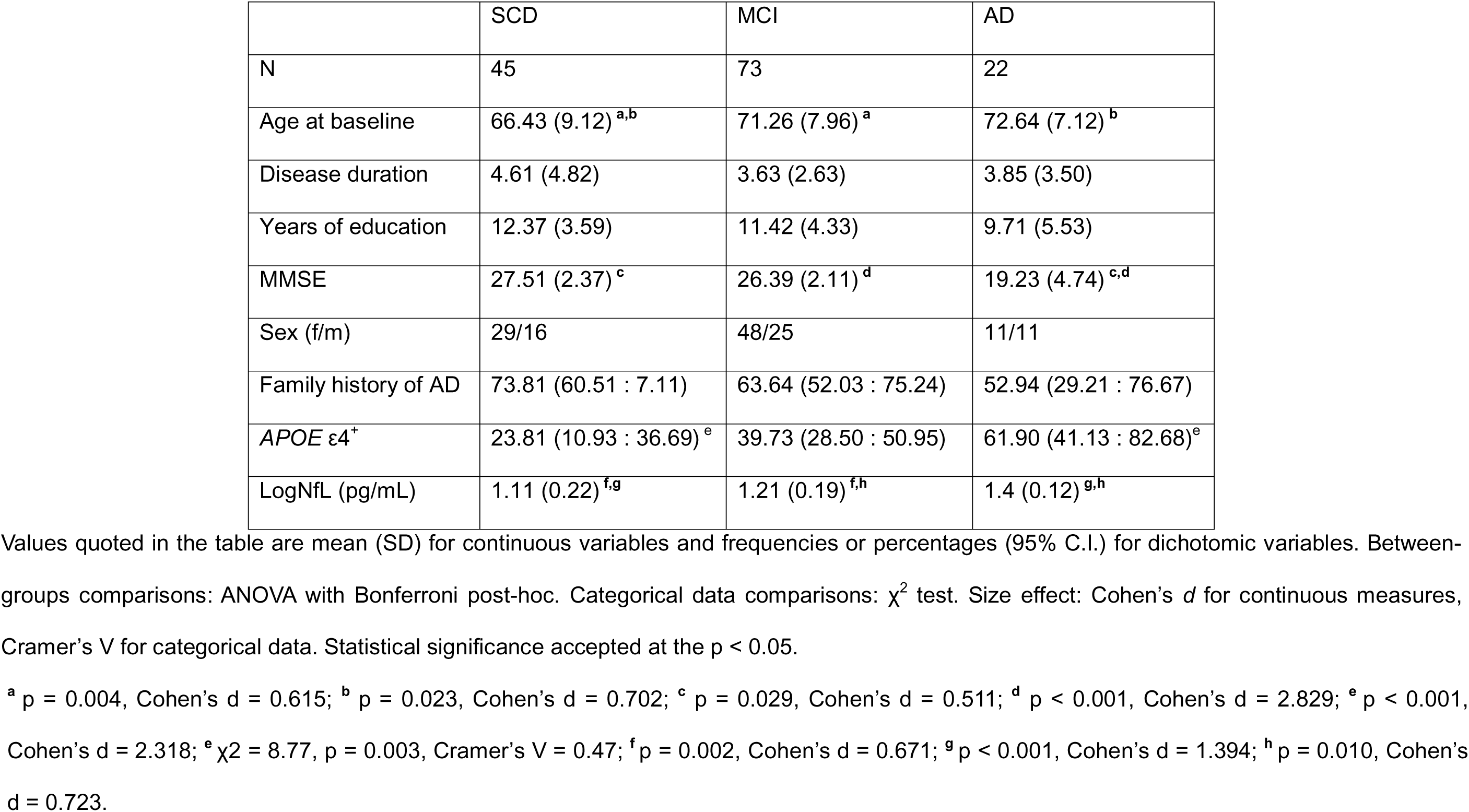
Comparisons between diagnosis groups

### 3.2. Biomarker profiles

Based on AD biomarker results, patients were classified as follows: 33 (30.0%) A-/T-N-, 11 (10.0%) A-/TN+, 12 (10.9%) A+, and 54 (49.1%) ATN+. Detailed distribution of CSF biomarker concentrations and frequencies of positive amyloid-PET and FDG-PET as well as comparisons between groups are reported in Supplementary table 2. Concordant and discordant results between CSF and PET are shown in Supplementary table 3.

### 3.3. Distribution of NfL concentration across ATN groups

NfL levels differed between ATN groups, also after adjusting for age, education, MMSE and *APOE* (*F*[3,96]=7.190, *p*<0.001, η^2^=0.160). Post-hoc analysis showed that ATN+ had higher NfL levels than A-/T-/N- (p<0.001, *d*=1.562), A-/TN+ (p=0.015, *d*=1.026) and A+ (p=0.001, *d*=1.419). There was no difference between A-/T-/N- and A-/TN+ (p=0.765, *d*=0.537), between A-/T-/N- and A+ (p=1.00, *d*=0.143) and between A-/TN+ and A+ (p=1.00, *d*=0.394) (Figure 1.B). Based on these results, for subsequent analysis, we merged A-/T-/N-, A-/TN+, and A+ into an ATN- group.

### 3.4. Distribution of NfL concentration across diagnosis/ATN subgroups

To explore the interaction between diagnosis and ATN group on NfL concentration, we classified patients according to diagnosis (SCD, MCI, AD-D) and ATN classification (ATN-, ATN+). As only one AD-D patient was ATN-, we did not split the AD-D group. The groups consisted of 21 SCD/ATN-, nine SCD/ATN+, 33 MCI/ATN-, 27 MCI/ATN+, and 20 AD-D. SCD/ATN-patients were younger than SCD/ATN+ (61.46±6.92 vs. 71.06 vs. 7.63, p=0.002). MCI/ATN-were younger (67.95±8.49 vs. 73.38±5.56, p=0.004) and had lower frequencies of *APOE*ε4+ (37.50% [95%C.I.=18.13:56.87] vs. 66.67% [95% C.I.=48.89:84.45], χ^2=^9.31, p=0.002) than MCI/ANT+. There were no differences in neuropsychological scores between SCD/ATN- and SCD/ATN+ and between MCI/ATN- and MCI/ATN+ (Supplementary table 4). NfL levels were significantly different between the diagnosis/ATN subgroups (*F*[5, 103]=13.50, *p*<0.001, η^2^=0.396). Differences in NfL concentration between groups were also confirmed after controlling for age, MMSE, MMSE and *APOE* (*F*[4,95]=6.95, p<0.001, partial η^2^=0.226, Supplementary table 1.B). Post-hoc analysis showed that SCD/ATN- had lower NfL concentrations than SCD/ATN+ (p=0.003, *d*=1.571), MCI/ATN+ (p<0.001, *d* =1.747) and AD-D (p<0.001, *d*=1.880). MCI/ATN- had lower NfL concentrations than MCI/ATN+ (p<0.001, *d*=1.343) and AD-D (p<0.001, *d*=1.476). There were no differences between SCD/ATN- and MCI/ATN- (p=1.00, *d*=0.447) or between SCD/ATN+, MCI/ATN+, and AD-D (p=1.00, *d*=0.133) (Figure 1.C).

### 3.5. Accuracy of plasma NfL in predicting ATN status

NfL showed good accuracy in distinguishing between ATN+ and ATN- in SCD and MCI groups (AUC 0.815 and 0.818, respectively). In the SCD group, a cut-off of 19.45 pg/mL yielded the maximum Youden Index and discriminated ATN- and ATN+ patients with excellent specificity (95.24 [95% C.I.=87.62:100]), good PPV and NPV (83.33% [95%C.I.=70.00:96.67]) but failed in sensitivity (55.56% [95%C.I.=37.77:73.34]). Similarly, in the MCI group a cut-off of 20.49 pg/mL had excellent specificity (93.94% [95%C.I.=87.90:99.98]), with very good PPV, fair NPV and poor sensitivity (62.96% [95%C.I.=50.74:75.18]). Finally, when we merged SCD and MCI, a cut-off of 20.03 pg/mL yielded the maximum Youden Index, discriminating ATN- and ATN+ patients with excellent specificity (94.44% [95%C.I.=89.71:99.18]), good PPV, fair NPV (78.46%[95%C.I.=69.97:86.95]) and poor sensitivity (61.1%[95%C.I.=51.04:71.18]) (Figure 2).

**Figure 2.**
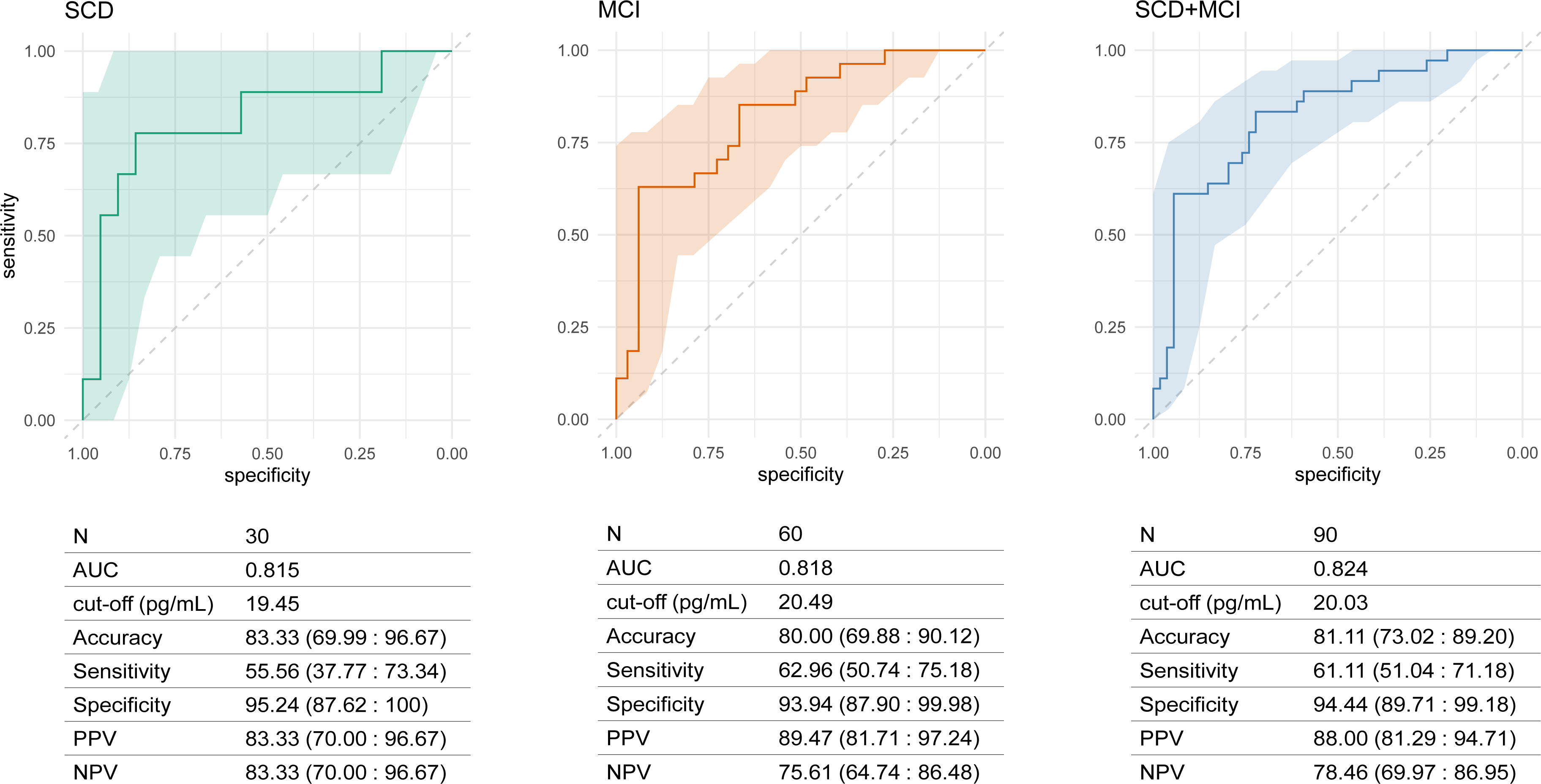
NfL accuracy in predicting ATN status. ROC curves for accuracy of NfL in distinguishing ATN- and ATN + groups in SCD, MCI and in SCD+MCI. Colored shapes indicate 95% C.I. Cut-off values estimated by Youden’s method. Accuracy, sensitivity, specificity, PPV and NPV are expressed as percentages (95% C.I.). AUC = area under the curve; PPV = positive predictive value; NPV = negative predictive value.

### 3.6. Trajectories of cognitive decline over time and comparison between progression groups

During follow-up, nine (30%) SCD patients progressed to MCI and were classified as p- SCD. Fourteen (29.79%) MCI patients developed dementia (p-MCI). None of the SCD patients developed dementia. Patients who did not progress were classified as np-SCD (21, 70.00%) and np-MCI (33, 70.21%). p-MCI had higher frequency of *APOE*ε4^+^ (p=0.017, *V*=0.348) and lower scores in minimental-state examination (p=0.002, *d*=1.239, Table 2) and in two tests for verbal memory (Supplementary table 5). There were no differences between np-SCD and p-SCD at baseline in demographic features, *APOE*ε4^+^, MMSE scores, or neuropsychological scoresTable 1. Comparisons between diagnosis groups

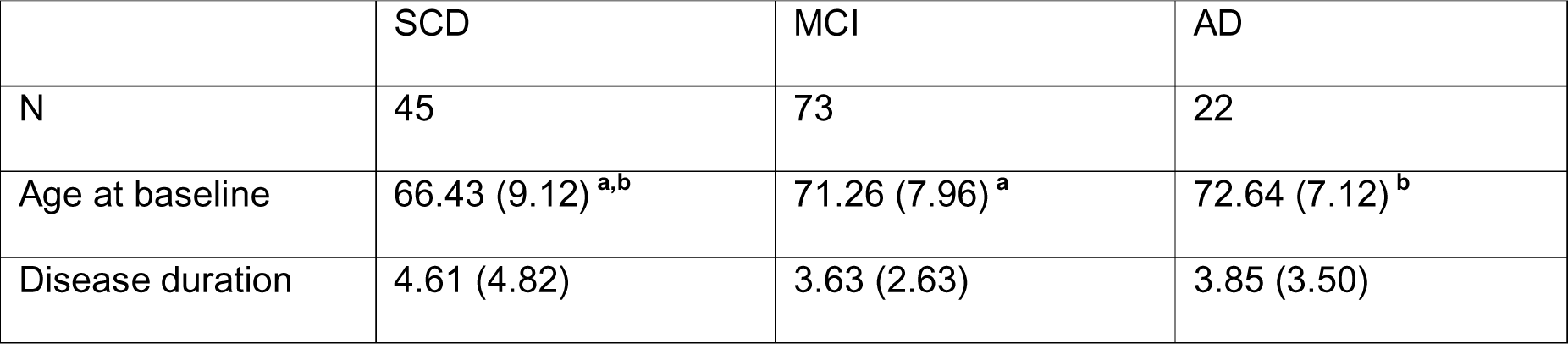
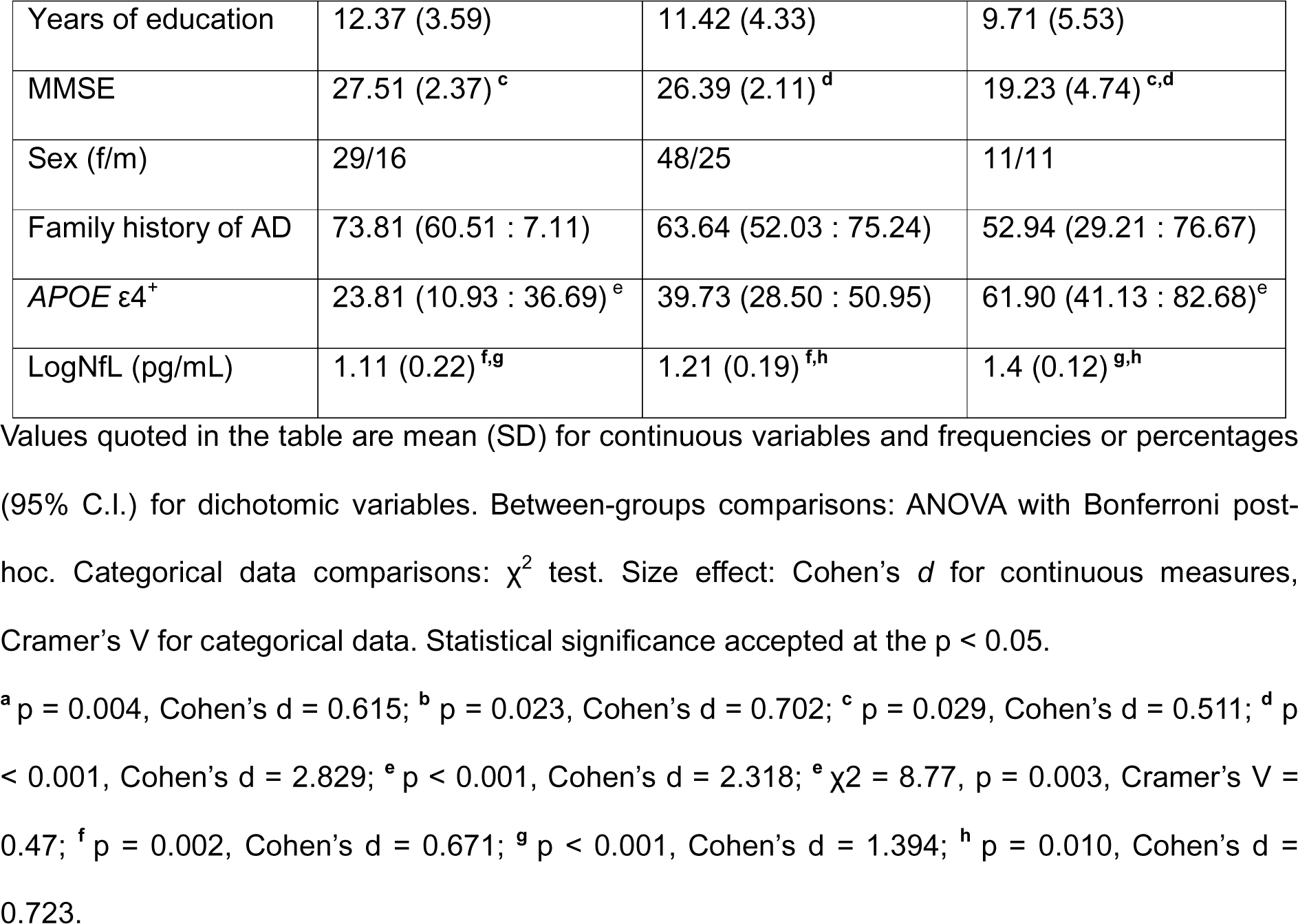

**Table 2.** Comparisons between progression groups

|  | np-SCD | p-SCD | np-MCI | p-MCI |
| --- | --- | --- | --- | --- |
| N | 21 | 9 | 33 | 14 |
| Age at baseline | 66.44 (6.60) | 68.20 (10.52) | 71.04 (7.62) | 73.57 (6.73) |
| Disease duration | 4.76 (5.97) | 4.24 (4.95) | 3.56 (2.47) | 2.84 (1.96) |
| Years of education | 12.29 (4.23) | 12.33 (3.32) | 10.97 (4.11) | 12.00 (4.09) |
| MMSE | 27.61 (4.42) <sup>a</sup> | 27.01 (2.42) <sup>b</sup> | 26.61 (1.86) <sup>c</sup> | 23.81 (3.05) <sup>a,b,c</sup> |
| Sex (f/m) | 12/9 | 7/2 | 21/12 | 10/4 |
| Family history of AD | 61.90 (41.13 : 82.68) | 77.78 (50.62 : 100) | 57.58 (40.71 : 74.44) | 35.71 (10.61 : 60.81) |
| *APOE* $\epsilon 4^+$ | 20.00 (2.47 : 37.53) <sup>†</sup> | 50.00 (15.35 : 84.65) | 27.27 (12.08 : 42.47) <sup>§</sup> | 64.29 (39.19 : 89.39) <sup>†§</sup> |
| LogNfL (pg/mL) | 1.10 (0.15) <sup>d</sup> | 1.23 (0.26) | 1.25 (0.19) <sup>e</sup> | 1.32 (0.16) <sup>d,e</sup> |

Values quoted in the table are mean (SD) for continuous variables and frequencies or percentages (95% C.I.) for dichotomic variables. Between-groups comparisons: ANOVA with Bonferroni post-hoc. Categorical data comparisons: $\chi^2$ test. Size effect: Cohen's d for continuous measures, Cramer's V for categorical data. Statistical significance accepted at the p < 0.05.
<sup>a</sup> p = 0.003, Cohen's d = 1.14; <sup>b</sup> p = 0.020, Cohen's d = 1.16; <sup>c</sup> p = 0.006, Cohen's d = 1.11; <sup>d</sup> p = 0.020, Cohen's d = 0.846; <sup>e</sup> p = 0.003, Cohen's d = 1.250. <sup>†</sup> p = 0.009, Cramer's V = 0.448; <sup>§</sup> p = 0.017, Cramer's V = 0.348.

Baseline NfL levels were significantly different between patients who progressed and patients who did not progress from SCD to MCI or from MCI to dementia (*F*[5,103]=5.06, p=0.003, η^2^=0.172). Post-hoc analysis showed that np-SCD had lower NfL concentration than np-MCI (p=0.020, *d*=0.846) and p-MCI groups (p=0.003, *d* =1.250) (Figure 1.D).

### 3.7. Effect of NfL group on the risk of progression of cognitive decline

We classified patients according to the previously identified cut-off values (NfL-=lower than cut-off values; NfL+=higher than cut-off value): seven (15.56%) SCD patients had NfL concentrations higher than 19.45 pg/mL and 23 (31.51%) MCI patients had NfL concentrations higher than 20.49 pg/mL. A Kaplan-Meier survival analysis showed a higher proportion of progression from SCD to MCI in the SCD/NfL+ group (80.00% [95%C.I.=44.94:100]) as compared to SCD/NfL-(22.73% [95%C.I.=5.22:40.24]; Log-rank χ^2^=9.79, p=0.002). There was no difference in the distribution of progression from MCI to AD dementia (Log-rank χ^2^=5.32, p=0.25) (Figure 3). To evaluate the effect of dichotomized NfL on the rate of conversion from SCD to MCI, adjusting for possible confounding factors, we performed Cox’s proportional hazards regression analysis considering progression time as time and including age at baseline, years of education and *APOE* genotype as covariates. The regression model was significant (χ^2^=9.702, p=0.002) and NfL group was the only significant variable (p=0.007, HR=7.10 [95%C.I.=1.71:29.50]).

**Figure 3.**
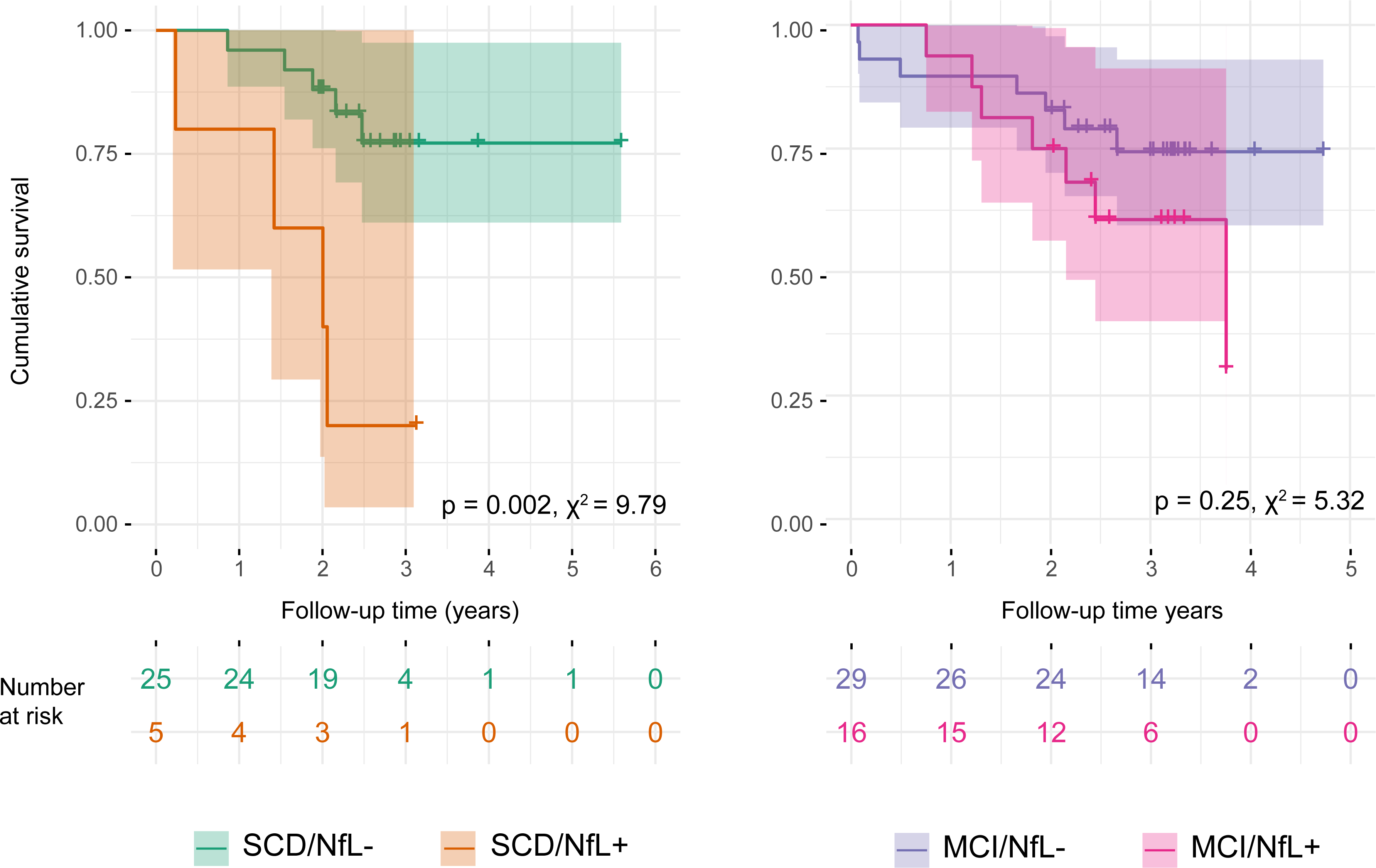
Kaplan–Meier survival analysis for comparison of distributions of progression from SCD to MCI and from MCI to AD between NfL- and NfL+ groups. For patients who progressed, follow-up time indicates the time of progression. Number at risk and p values for pairwise log-rank comparisons between groups are reported. Colored shapes indicate 95% C.I.

### 3.8. Accuracy of plasma NfL in predicting progression of cognitive decline

In the SCD group, the cut-off of 19.45 pg/mL showed good accuracy (80.00% [95%C.I.=65.69:94.31]) with excellent specificity (95.24% [95%C.I.=87.62:100]), good PPV and NPV (80.00% [95%C.I.=65.69:94.31]), but not acceptable sensitivity (44.44% (95%C.I.=26.66:62.23]) in predicting progression to MCI. In the MCI group, NfL, with the cut-off value of 20.49 pg/mL, had fair specificity (70.97% [95%C.I.=57.71:84.23]) and NPV (75.86% [95%C.I.=63.63-:88.10]), but not acceptable sensitivity and PPV (≤50%) in predicting progression to dementia. Finally, considering SCD and MCI together, NfL had fair specificity and NPV (77.78% [95%C.I.=68.49:87.06]) but not acceptable sensitivity and PPV (47.83% [95%C.I.=36.67:58.98]) (Table 4).

**Table 3.**
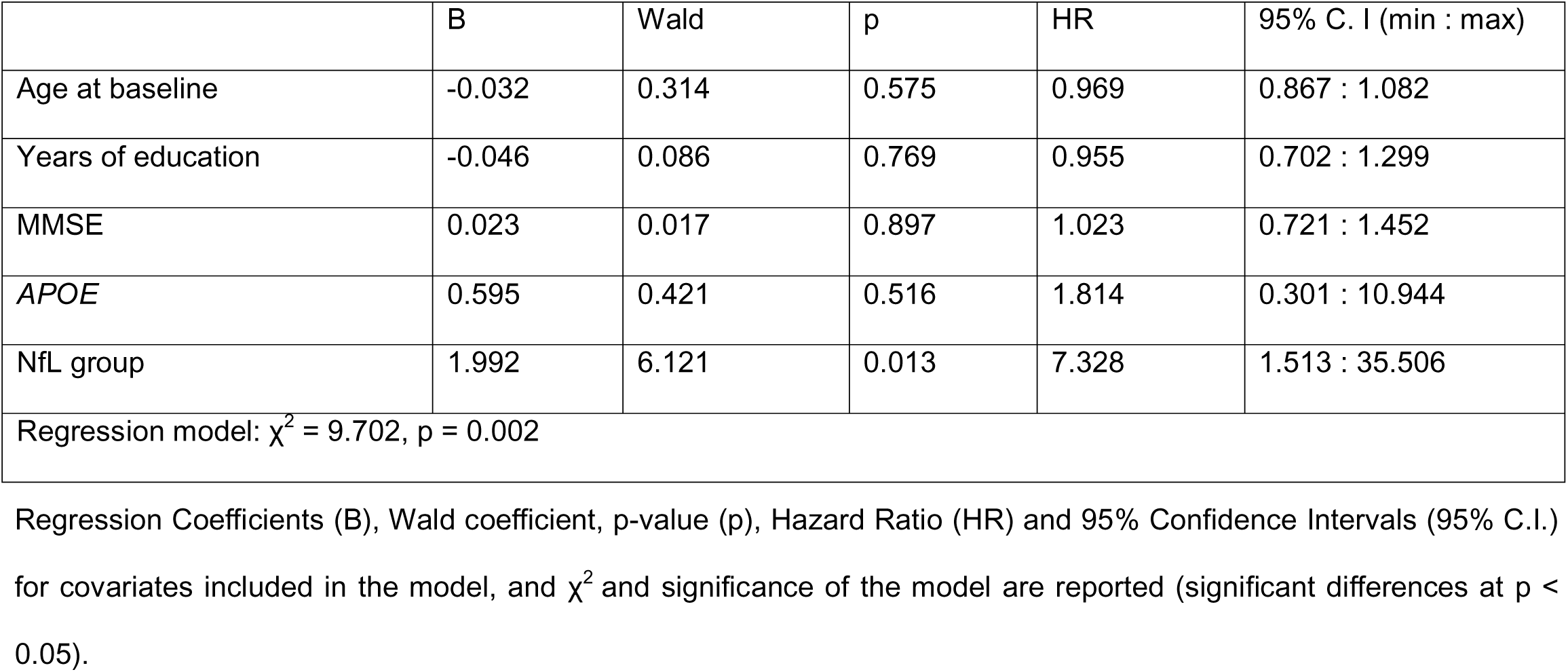
Cox’s proportional hazards regression analysis

**Table 4.** Accuracy of NfL concentration at baseline and rate of change in predicting the progression of cognitive decline

|  | SCD | MCI | SCD+MCI | Rate of change (MCI) |
| --- | --- | --- | --- | --- |
| N | 30 | 47 | 77 | 29 |
| cut-off (pg/mL) | 19.45 | 20.45 | 20.03 | 1.64 |
| Accuracy | 80.00 (65.69 : 94.31) | 64.44 (50.46 : 78.43) | 68.83 (58.49 : 79.18) | 92. 86 (83.32 : 100) |
| Sensitivity | 44.44 (26.66 : 62.23) | 50.00 (35.39 : 64.61) | 47.83 (36.67 : 58.98) | 100.00 |
| Specificity | 95.24 (87.62 : 100) | 70.97 (57.71 : 84.23) | 77.78 (68.49 : 87.06) | 91.30 (80.87 : 10) |
| PPV | 80.00 (65.69 : 94.31) | 43.75 (29.26 : 58.24) | 47.83 (36.67 : 58.98) | 71.43 (54.70 : 88.16) |
| NPV | 80.00 (65.69 : 94.31) | 75.86 (63.63 : 88.10) | 77.78 (68.49 : 87.06) | 100 |

Cut-off values estimated by Youden's method. Accuracy, sensitivity, specificity, PPV and NPV are expressed as percentages (95% C.I.). AUC = area under the curve; PPV = positive predictive value; NPV = negative predictive value.

### 3.9. Change of NfL concentration over time

Forty-eight patients (19 SCD, 29 MCI) underwent new blood collection for NfL measurement two years (T2) after baseline collection (T1). NfL measures were highly consistent over time (ICC=0.84 [95%C.I.=0.73:0.91], p<0.001). Considering the whole sample, the mean NfL change (ΔNfL) was 1.13±5.47 pg/mL in two years (0.71±2.98 pg/mL per year), with no differences between SCD and MCI. ΔNfL was correlated with age at baseline (Pearson=0.341, p=0.017), while there was no effect of disease duration, education, sex, family history of AD dementia, and *APOE* genotype on NfL change.

### 3.10. Effect of ATN status and progression of cognitive decline on NfL over time

A repeated measures ANOVA showed a significant interaction between change in NfL and progression of cognitive decline (*F* [3,44]= 5.2, *p*=0.032, η^2^=0.014), confirmed also after age-adjustment (*F*[3,41]=4.28, p=0.010, η^2^=0.239, Supplementary table 1.C). Post-hoc analysis showed that this effect was significant between np-SCD and p-MCI (t=-4.32, p<0.001) and between np-MCI and p-MCI (t=-2.93, p=0.033, Figure 4.A). In particular, NfL concentration showed a change of 1.63±2.50 pg/mL (0.81±1.25 pg/mL per year) and of - 1.39±3.88 pg/mL (−0.13±3.24 pg/mL per year) in np-SCD and np-MCI respectively, and an increase of 7.05±8.12 pg/mL (3.52±4.06 pg/mL per year) in p-MCI. The effect of ATN status on NfL change did not reach significance (*F*[1,31]=2.80, p=0.056, η^2^=0.023). Nevertheless, when performing a post-hoc analysis considering the ATN groups (A-/T-/N-, A-/TN+, A+, and ATN+), we found a different effect on NfL change between A+ and ATN+ (t=-3.15, p=0.024) (Figure 4.B). In particular, NfL showed a decrease of −1.94±3.32 pg/mL (−0.97±1.66 pg/mL per year) in the A+ group and an increase of 3.07±7.21 pg/mL (1.53±3.60 pg/mL per year) in the ATN+.

**Figure 4.**
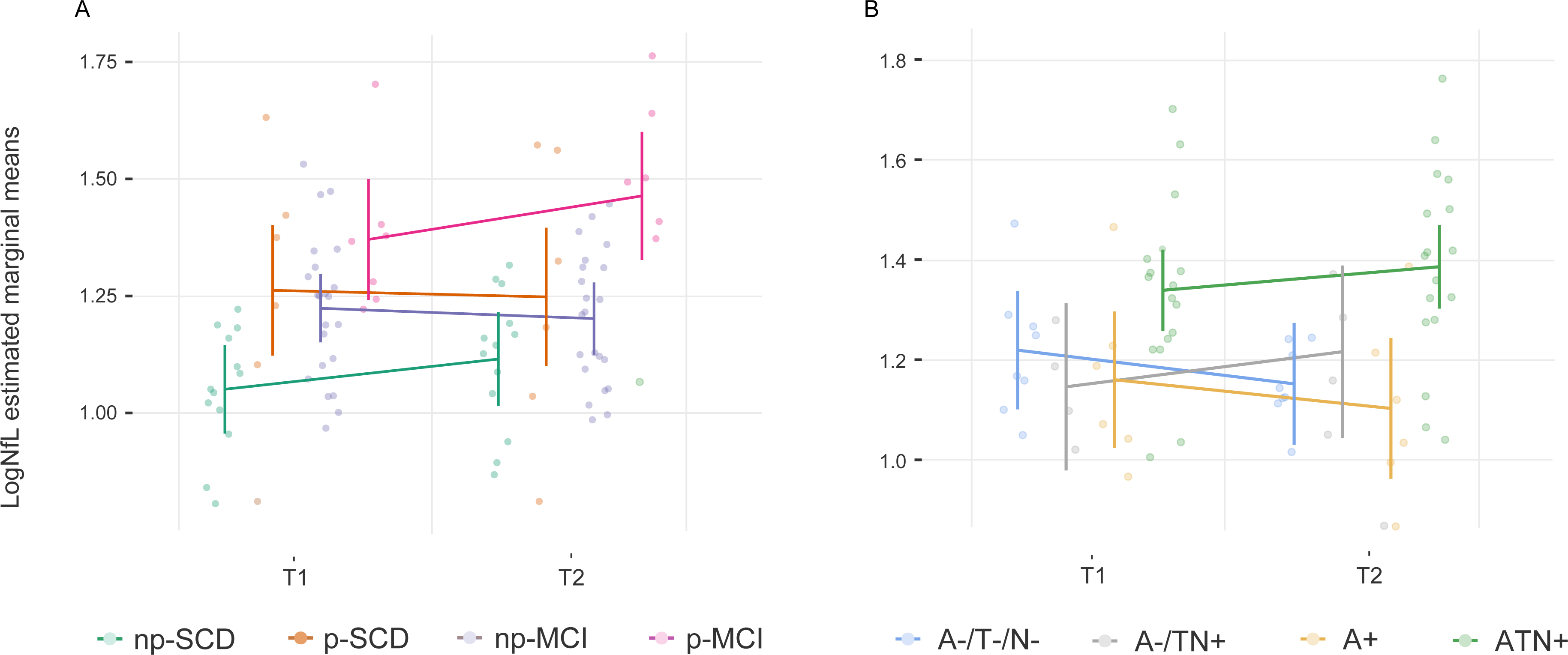
Change in logNfL distribution by progression and ATN groups. T1 and T2 indicate the first and second blood collection for plasma NfL measurement. A. Effect of cognitive decline progression on LogNfL change: *F* [3, 44] = 5.2, *p* = 0.032, η^2^ = 0.014. Post-hoc analysis: - np-SCD vs. p-SCD (mean difference = −0.18, p = 0.214) - np-SCD vs. np-MCI (mean difference = −0.13, p = 0.223) - np-SCD vs. p-MCI (mean difference = −0.34, p < 0.001) - p-SCD vs. np-MCI (mean difference = 0.05, p = 1.00) - p-SCD vs. p-MCI (mean difference = −0.16, p = 1.00) - np-MCI vs. p-MCI (mean difference = −0.21, p = 0.033) B. Effect of ATN on NfL change: *F* [1, 31]= 2.80, *p* = 0.056, η^2^ = 0.213). Post-hoc analysis: - A-/T-/N- vs. A-/TN+ (mean difference = 0.00, p = 1.00) - A-/T-/N- vs. A+ (mean difference = 0.05, p = 1.00) - A-/T-/N- vs. ATN+ (mean difference = −0.18, p = 0.077) - A-/TN+ vs. A+ (mean difference = 0.05, p = 1.00) - A-/TN+ vs. ATN+ (mean difference = −0.18, p = 0.269) - A+ vs. ATN + (mean difference = −0.23, p = 0.024)

### 3.11. Diagnostic accuracy of NfL change rate in predicting the progression from MCI to AD dementia

We tested the accuracy of NfL change rate (ΔNfL/per year) in distinguishing between np-MCI an p-MCI: a cut-off of 1.64 pg/mL per year showed the highest Youden index, providing very good accuracy (92.86% [95%C.I.=83.32:100], AUC=0.954) with very high sensitivity (100%), specificity (91.30% [C.I.95%=80.87:100]) and NPV (100%) in distinguishing between np-MCI and p-MCI. Also in this case, PPV was fair (71.43% [95%C.I.=54.70:88.16]). We did not perform the same analysis in the SCD group, as the repeated measures ANOVA did not show a significant effect of progression on NfL change in this group.

## 4. Discussion

We showed that SCD and MCI patients with biomarkers consistent with AD had higher plasma NfL concentration than patients with normal biomarkers or with demonstration of isolated Aβ pathology. We identified cut-off values to distinguish ATN- and ATN+ with very good accuracy (19.45 and 20.49 pg/mL in SCD and MCI respectively). Interestingly, these cut-off values were very close to the cut-off value (20 pg/mL) identified by Simrén et al. in a large cohort of healthy individuals^23^.

Moreover, NfL concentration in SCD and MCI patients in the ATN+ group did not differed from patients with dementia due to AD. This finding is particularly interesting for SCD patients and is in line with longitudinal studies showing that blood NfL levels increase more than a decade before the onset of clinical manifestations in carriers of amyloid precursor protein (*APP*), presenilin 1 (*PSEN1*), and presenilin 2 (*PSEN2*) mutations^24^.

We then tested whether the identified cut-off values were also able to detect progression from SCD to MCI and from MCI to AD. In this case, NfL showed high prognostic performance in the SCD group, being able to predict and exclude progression to MCI over two years, with 80% PPV and NPV. Finally, we explored the change in NfL concentrations over time. Overall, NfL concentration increased by 0.71 pg/mL per year, in line with previous reports^10^. Nevertheless, we showed that the rate of increase was higher in patients with ATN+ status than in patients with isolated A+ and in patients who progressed from MCI to AD dementia compared to patients who did not progress. In particular, the rate of increase in MCI patients who progressed to dementia was approximately 3.5 times higher than that in MCI patients who remained stable, which is consistent with a previous result by de Wolf et al.^15^. Moreover, we showed that an increase lower than 1.64 pg/mL per year can exclude progression from MCI to AD with high accuracy, in line with a previous study that showed that the rate of change of serum NfL was able to discriminate carriers of a mutation in *APP*, *PSEN1* or *PSEN2* genes from non-carriers^14^.

The high specificity of NfL in distinguishing between ATN+ and ATN- and between SCD patients who progressed and those who did not progress to MCI might appear in contrast with the general assumption that NfL is a highly sensitive but poorly specific biomarker^10^. Indeed, previous studies showed low accuracy of NfL when distinguishing AD from other neurological diseases^25^. In contrast, when compared to cognitively healthy individuals, NfL was shown to be the most accurate blood-based biomarker^25^. Therefore, our results may suggest that, if applied to unimpaired patients complaining of memory decline (in which other possible non-degenerative causes have been ruled out by first-line assessments), plasma NfL may be highly suggestive of underlying AD.

It should be noted that patients with isolated Aβ biomarker positivity had the same NfL levels as patients with normal AD biomarkers, in line with previous reports^12, 13, 26^ and with the hypothesis raised by Benet et al.^27^ that NfL concentration increases when Aβ pathology and tauopathy are associated. This evidence may have a relevant clinical implications in terms of the risk of AD and progression to dementia. Indeed, although being part of the AD continuum, isolated Aβ pathology is not sufficient to define AD^22^ and is associated with the lowest risk of AD dementia^28^.

Our study has some limitations: 1) the relatively small number of patients, particularly after splitting into subgroups; 2) the follow-up time was fairly short; 3) not all the patients underwent NfL assessment at follow-up; 4) we did not include a sample of healthy controls; 5) tau pathology was assessed only by CSF p-tau, with a possible underestimation of positive T frequencies.

On the other hand, Aβ was assessed by both CSF Aβ42 and Aβ42/40 and amyloid-PET, as well as neurodegeneration was assessed by CSF total-tau and FDG-PET. This is the first strength of our study. Second, this is one of the few studies^28–30^ that tested plasma NfL in a SCD cohort. Notably, while most studies assessed the accuracy of NfL in detecting Aβ pathology^12, 13^, we classified patients according to ATN status, also considering tau pathology and neurodegeneration biomarkers. Moreover, follow-up data were used to validate the performance of the identified cut-off value also in predicting progression to MCI and dementia. Finally, our study explores one of the main advantages of blood-based biomarkers: as they are non-invasive, their measurement can be repeated many times. In this sense, our study adds useful information showing that the NfL trajectory may accurately distinguish between patients who will progress to dementia and patients who will not.

In conclusion, our results have potential implications for the clinical management of patients with SCD and MCI. Individuals with negative NfL levels had a lower risk of carrying AD and progressing to MCI or dementia. Therefore, they may require monitoring for cognitive decline and investigation of other causes of cognitive decline. In contrast, patients with NfL levels exceeding the cut-off value had a higher risk of AD and cognitive decline progression. In the era of DMTs, this may enable earlier identification of patients suitable for treatment in the earliest stages of the disease.

## Data Availability

All data produced in the present study are available upon reasonable request to the authors

## Notes

**Funding:** This project was funded by Tuscany Region - PRedicting the EVolution of SubjectIvE Cognitive Decline to Alzheimer’s Disease With machine learning - PREVIEW - CUP. D18D20001300002

### Competing Interest Statement

The authors have declared no competing interest.

### Clinical Protocols

https://clinicaltrials.gov/ct2/show/NCT05569083

### Funding Statement

This project was funded by Tuscany Region - PRedicting the EVolution of SubjectIvE Cognitive Decline to Alzheimer's Disease With machine learning - PREVIEW - CUP. D18D20001300002

