## Supplementary tables for "Plasma neurofilament light chain predicts Alzheimer’s disease in patients with subjective cognitive decline and mild cognitive impairment: a longitudinal study": Supplementary tables.pdf

Supplementary table 1. Covariate adjusted models

| A. Analysis of covariance for NfL by diagnosis groups (SCD, MCI, AD) | | | | |
|---|---|---|---|---|
| | df | F | p | partial $\eta^2$ |
| Adjusted model | 6 | 12.21 | <0.001 | 0.381 |
| Age at baseline | 1 | 40.985 | <0.001 | 0.256 |
| MMSE | 1 | 0.647 | 0.423 | 0.005 |
| *APOE* | 1 | 0.013 | 0.910 | <0.001 |
| Diagnosis groups | 2 | 30.505 | 0.033 | 0.056 |
| Error | 119 | | | |
| Adjusted $R^2$ = 0.350 | | | | |
| B. Analysis of covariance for NfL by diagnosis/ATN groups | | | | |
| | df | F | p | partial $\eta^2$ |
| Adjusted model | 1 | 10.68 | <0.001 | 0.473 |
| Age at baseline | 1 | 13.39 | <0.001 | 0.124 |
| MMSE | 1 | 0.03 | 0.860 | 0.000 |
| *APOE* | 1 | 2.08 | 0.153 | 0.021 |
| Diagnosis/ATN groups | 4 | 6.95 | <0.001 | 0.226 |
| Error | 95 | | | |
| Adjusted $R^2$ = 0.429 | | | | |
| C. Analysis of covariance for NfL change by progression groups | | | | |
| | df | F | p | partial $\eta^2$ |
| NfL change | 2 | 5.87 | 0.020 | 0.125 |
| NfL change*age at baseline | 1 | 6.78 | p = 0.013 | 0.142 |
| NfL change*progression | 3 | 4.28 | p = 0.010 | 0.239 |
| Error | 41 | | | |

df = degrees of freedom

Supplementary table 2. Comparison of biomarker concentration and prevalence between groups

| | SCD | MCI | AD |
|---|---|---|---|
| CSF | | | |
| N | 30 | 60 | 20 |
| $A\beta_{42}$ (pg/mL) | 1066.06 (400.61) [a] | 929.21 (450.78) [b] | 518.33 (146.6) [a,b] |
| $A\beta_{42}/A\beta_{40}$, | 0.08 (0.02) [c] | 0.07 (0.02) [d] | 0.04 (0.01) [c,d] |
| p-tau (pg/mL) | 55.00 (31.54) [e] | 68.48 (50.89) [f] | 129.53 (60.8) [e,f] |
| t-tau (pg/mL) | 417.35 (221.31) [g] | 456 (296.9) [h] | 784.38 (349.15) [g,h] |
| Neuroimaging | | | |
| Amyloid-PET + | 9/16 (56.25) | 6/9 (66.67) | 2/3 (66.67) |
| FDG-PET + | 7/23 (30.43) [^,+] | 18/51 (35.29)[+] | 17/19 (89.47) [^,+] |
| ATN | | | |
| A+ | 13/30 (43.33)[ʃ] | 33/60 (55.00)[°] | 20/20 (100)[ʃ °] |
| T+ | 9/30 (30.00)^ | 30/60 (50.00)[¶] | 19/20 (95.00) [^¶] |
| N+ | 11/30 (36.67)[†] | 34/60 (56.67)[§] | 19/20 (95.00) [† §] |

Values quoted in the table are mean (SD) for continuous variables and frequencies of positive on total number (percentages) for dichotomic variables. Statistical significance accepted at the $p < 0.05$.

[a] $p < 0.001$, d = 1.05 [b] $p < 0.001$, d = 2.21; [c] $p < 0.001$, d = 2.01; [d] $p < 0.001$, d = 1.74; [e] $p < 0.001$, Cohen's d = 1.64; [f] $p < 0.001$, Cohen's d = 1.17; [g] $p < 0.001$ Cohen's d = 1.45; [h] $p < 0.001$, Cohen's d = 1.13.

^$\chi^2$ = 14.8, $p < 0.001$, V = 0.60; [+] $\chi^2$ = 16.3, $p < 0.001$, V = 0.48; [ʃ]$\chi^2$ = 19.41, $p < 0.001$, V = 0.59; [°] $\chi^2$ = 14.91, $p < 0.001$, V = 0.42; ^$\chi^2$ = 14.50, $p < 0.001$, V = 0.54; [¶] $\chi^2$ = 7.58, p = 0.006, V = 0.31; [†]$\chi^2$ = 17.00, $p < 0.001$, V = 0.58; [§] $\chi^2$ = 9.86, p = 0.002, V = 0.31.

Supplementary table 3. Convergence results of AD biomarkers

| A | Amyloid-PET - | Amyloid-PET + |
|---|---|---|
| CSF A- | 8 (5 SCD, 3 MCI) | 3 (2 SCD, 1 MCI) |
| CSF A + | 3 (2 SCD, 1 AD) | 14 (7 SCD, 5 MCI, 2 AD) |
| B | FDG-PET − | FDG-PET + |
| CSF N − | 32 (11 SCD, 21 MCI) | 3 (2 SCD, 1 MCI) |
| CSF N + | 19 (5 SCD, 12, MCI, 2 AD) | 39 (5 SCD, 17 MCI, 17 AD) |

CSF A- : patients with both negative CSF $A\beta_{42}$ and $A\beta_{42}/A\beta_{40}$

CSF A+: patients with positive CSF $A\beta_{42}/A\beta_{40}$ and/or $A\beta_{42}/A\beta_{40}$

CSF N-: patients with negative CSF t-tau

CSF N+: patients with positive CSF t-tau

Supplementary table 4. Comparison in neuropsychological scores between diagnosis/ATN groups

| | SCD/ATN- | SCD/ATN+ | MCI/ATN- | MCI/ATN+ |
|---|---|---|---|---|
| N | 21 | 9 | 33 | 27 |
| RAVLT immediate recall | 49.29 (6.96) | 47.46 (6.52) | 40.04 (9.67) | 33.93 (9.41) |
| Short story immediate recall | 12.76 (3.36) | 8.25 (2.06) | 7.76 (4.06) | 7.38 (3.62) |
| RAVT delayed recall | 10.52 (2.45) | 10.94 (3.55) | 7.02 (4.31) | 4.64 (3.82) |
| Short story delayed recall | 16.71 (3.77) | 13.00 (5.35) | 11.10 (4.13) | 7.46 (7.17) |
| Rey-Osterrieth complex figure recall | 21.31 (4.06) | 19.45 (5.42) | 14.16 (6.03) | 11.31 (7.90) |
| Phonemic Fluency Task | 38.12 (8.61) | 42.60 (13.18) | 34.24 (12.10) | 33.04 (9.74) |
| Category Fluency Task | 46.00 (10.16) | 43.00 (11.05) | 38.66 (12.036) | 31.96 (7.66) |
| Attentional matrices | 51.60 (5.54) | 47.00 (10.64) | 47.99 (8.40) | 46.50 (10.23) |
| Trail Making Test A | 25.72 (12.76) | 31.40 (20.07) | 64.10 (105.92) | 68.00 (156.01) |
| Trail Making Test B | 56.33 (44.22) | 79.00 (55.68) | 219.47 (227.88) | 253.43 (259.19) |
| Trail Making Test B-A | 30.33 (35.51) | 47.80 (38.90) | 155.37 (174.37) | 185.87 (242.34) |
| Stroop Test | 17.22 (7.40) | 18.05 (17.07) | 56.67 (145.45) | 28.61 (22.00) |
| Digit span | 6.41 (0.80) | 5.75 (1.26) | 5.64 (1.29) | 5.64 (0.84) |
| Visuo-spatial Span | 4.94 (0.77) | 5.25 (0.50) | 4.59 (0.80) | 5.07 (1.00) |
| Rey-Osterrieth complex figure recall | 34.00 (1.82) | 33.60 (3.46) | 31.37 (2.82) | 32.13 (4.24) |

Values quoted in the table are mean (SD). RAVLT = Rey Auditory Verbal Learning Test

Supplementary table 5. Comparison in neuropsychological scores between progression groups

| | np-SCD | p-SCD | np-MCI | p-MCI |
|---|---|---|---|---|
| N | 21 | 9 | 33 | 14 |
| RAVLT immediate recall | 49.83 (5.79) | 50.72 (3.88) | 38.96 (7.53) | 27.48 (7.40) |
| Short story immediate recall | 12.18 (3.25) | 8.50 (1.73) | 9.17 (3.41) | 3.20 (3.35) |
| RAVT delayed recall | 10.58 (2.94) | 9.84 (2.68) | 6.53 (3.81) [a] | 2.17 (2.78) [a] |
| Short story delayed recall | 15.64 (3.01) | 13.00 (4.69) | 12.08 (6.04) [b] | 1.80 (3.49) [b] |
| Rey-Osterrieth complex figure recall | 22.29 (4.45) | 16.75 (3.35) | 14.26 (8.03) | 8.50 (4.88) |
| Phonemic Fluency Task | 39.73 (11.95) | 38.80 (6.38) | 36.67 (9.81) | 31.50 (12.33) |
| Category Fluency Task | 46.30 (12.22) | 41.80 (8.59) | 37.00 (10.82) | 31.58 (10.31) |
| Attentional matrices | 51.50 (6.06) | 53.94 (2.24) | 49.30 (6.85) | 47.75 (9.45) |
| Trail Making Test A | 27.83 (12.52) | 29.60 (18.02) | 42.00 (49.31) | 34.67 (16.15) |
| Trail Making Test B | 65.33 (51.05) | 82.80 (53.19) | 202.32 (217.13) | 226.00 (237.73) |
| Trail Making Test B-A | 37.50 (42.13) | 53.60 (41.11) | 160.55 (195.36) | 191.75 (230.40) |
| Stroop Test | 15.042 (7.65) | 18.55 (15.55) | 23.55 (18.28) | 97.80 (231.01) |
| Digit span | 5.73 (0.91) | 6.50 (1.00) | 5.62 (0.96) | 5.60 (1.82) |
| Visuo-spatial Span | 4.70 (0.95) | 5.50 (1.00) | 4.62 (0.96) | 5.20 (0.84) |
| Rey-Osterrieth complex figure recall | 33.77 (1.93) | 33.30 (3.21) | 31.85 (3.65) | 32.64 (2.84) |

Values quoted in the table are mean (SD). Between-groups comparisons: t-test (np-SCD vs. p-SCD and np-MCI vs. p-MCI). RAVLT = Rey Auditory Verbal Learning Test.

[a] $t = 3.48$, $p = 0.001$; [b] $t = 3.52$, $p = 0.003$
